# Early Life Determinants of Forward Compression Wave Intensity in Adults

**DOI:** 10.64898/2026.05.26.26354176

**Authors:** Andrew Haynes, Jonathan P. Mynard, Mart van der Veen, Jennie Carson, Daniel J. Green

**Author notes:** **Corresponding author:** Daniel J Green. The University of Western Australia M408, Nedlands WA 6009, Australia.

## Abstract

**Intro:** Characteristics of the pulse wave transmitted through the carotid arteries are predictive of cognitive decline and cerebrovascular health in humans. This study aimed to identify risk factor trajectories in childhood, adolescence and early adulthood that are associated with forward compression wave intensity (FCWI) in the common carotid artery in adults aged 28 years.

**Methods:** Systolic blood pressure (SBP), body mass index (BMI) and fasting blood glucose (FBG) measured at multiple time-points when participants were aged between 8-20 years were included in a trajectory analysis. At age 28 years, FCWI was measured in 402 (M=206, F=196) participants who underwent a Duplex ultrasound assessment of the common carotid artery. Statistical analysis assessed differences in FCWI between each trajectory group for males and females separately.

**Results:** In males, four trajectory groups were identified for BMI, three for SBP, and two for FBG. In females, three trajectory groups were identified for BMI, SBP, and FG. In males, having higher BMI (*P*=0.006), SBP (*P*=0.021) and FBG (*P*=0.002) from ages 8-20 years was associated with greater FCWI at age 28 years. In females, no associations were found between FCWI at age 28-years and trajectory groups for BMI (*P*=0.185), SBP (*P*=0.289) or FBG (*P*=0.070).

**Conclusion:** Having high BMI, SBP and FBG throughout childhood, adolescence and early adulthood was associated with higher FCWI in the carotid artery at age 28 years in males, but not females. This may have a direct impact on the etiology of cognitive decline and cerebrovascular disease in later life.

## Introduction

The ageing brain is susceptible to a variety of neurodegenerative disorders that impact cognition (e.g. Alzheimer’s disease, dementia), and these often occur alongside the development of cerebrovascular disease. Worldwide, over 12 million people experience a stroke per annum (1), and ∼10 million are diagnosed with dementia (2). The etiology of these conditions is multi-factorial but includes the lifelong impact of exposure to modifiable risk factors (1–3).

The hemodynamic impact of the blood pressure/flow pulse wave on the cerebral vascular beds has emerged as a potential contributor to the pathogenesis of cognitive and cerebrovascular pathology (4). Approximately 85% of the blood-brain barrier is made up of capillaries, and the single layer of endothelial cells that make up the capillary walls are particularly vulnerable to damage in response to high hemodynamic stresses (5). Studies examining links between hemodynamic forces and small vessel dysfunction and damage utilized pulse-pressure and pulse-wave velocity to estimate mechanical stress on the vessel walls (6, 7). However, more recent studies have introduced carotid pulse wave analysis, and specifically forward compression wave intensity (FCWI) that represents the energy of the advancing pulse wavefront (4, 8). Indeed, there is evidence to suggest that peak FCWI may predict cognitive decline in older adults (4).

Prevention of age-related diseases affecting cognition and cerebrovascular health should logically address early life antecedents that increase susceptibility. There is an established and growing literature highlighting the relationship between risk factor exposure in childhood, adolescence, and early adulthood, on chronic diseases that manifest in older adulthood (9, 10). Identifying early life determinants of adult disease will strengthen the rationale for early disease detection and direct targeted prevention strategies. In studies that measure outcomes sequentially in the same participants (i.e. cohort studies), trajectory analysis provides a powerful statistical approach that may be superior to treating data at each timepoint independently (11). Trajectory analysis has previously been conducted in the Raine Study, a birth cohort study based in Perth, Western Australia, in which participants have attended multiple follow-ups throughout their life (12–14).

This study aimed to determine whether risk factor trajectories (body mass index BMI, systolic blood pressure SBP, and fasting blood glucose FBG) in the Raine cohort were associated with FCWI in the carotid artery at age 28 years. We hypothesized that a higher risk factor profile across childhood, adolescence and early-adulthood would be associated with greater FCWI in adulthood.

## Methods

Participants were Generation 2 (Gen2) of the Raine Study, whose pregnant mothers were recruited between May 1989 and November 1991 in Perth, Western Australia. The 2,868 babies entered into the study subsequently attended regular follow-ups throughout childhood and adolescence (12).

### Written Informed Consent

This study conformed to the Declaration of Helsinki and was approved by The University of Western Australia Human Research Ethics Committee. Written, informed consent was obtained at all time-points, initially from the parents until the children reached 17 years, and thereafter provided their own consent.

### Risk Factor Trajectory Modelling

Participants attended follow-up appointments at regular timepoints throughout their life (∼28 years at latest follow-up); and some of these data were used to develop risk factor trajectories for BMI (z-scores), resting blood pressure and fasting blood glucose, using latent class modelling. Measurements of BMI and SBP took place at 5 follow-ups over a 12-year period when participants were aged 8, 10, 14, 17 and 20 years. Measurement of FBG took place at 4 follow-ups when participants were aged 8, 14, 17 and 20 years (i.e. blood tests were not included at the 10-year follow-up), and these were included in FBG trajectory development. Due to the known differences in risk factors such as blood pressure and BMI (15, 16) between males and females, trajectories were developed for males and females separately, an approach we have used in the past with this cohort (13). Group-based trajectory modelling was used to identify developmental patterns in BMI (z-scores), SBP and FBG using a censored normal distribution with the trajectory plugin in Stata 18.0 for Windows (StataCorp, College Station, TX) (17). SBP trajectories were adjusted for height, and BMI z-scores were calculated using 2022 growth charts developed by the Centre for Chronic Disease and Health Promotion. Fasting glucose values were treated as continuous outcomes without transformation. Participants were included if they had data for at least three timepoints out of the 5 follow-ups, and also participated in the 28-year follow-up which served as the outcome time point (details below). Model selection was guided by the Bayesian Information Criterion (BIC), average posterior probabilities ≥0.80, odds of correct classification > 5, the proportion of individuals assigned to each group, and the interpretability of trajectory shapes (e.g., “low healthy,” “healthy,” “high healthy”). These criteria ensured well-fitting, distinct, and clinically relevant trajectory groups.

### Forward Compression Wave Intensity

Between March 2018 and December 2019, when participants were approximately 28 years of age (Gen2-28), 1,997 participants were contacted (in order of birth oldest – youngest) with an invitation to attend a follow-up appointment. Participants arrived at the laboratory in the morning between 7-9 am, following an overnight fast, having not consumed any liquid other than water and not engaged in exercise the morning of the appointment. Participants accepting the invitation, received a duplex ultrasound assessment of the common carotid artery (CCA), in which artery diameter and blood velocity across the cardiac cycle were measured. Prior to having the ultrasound scan, participants lay in a supine position in a temperature-controlled room for at least 15 minutes. A portable high resolution ultrasound machine (T3200; Terason, Burlington, MA) with 12 MHz linear array transducer was used to obtain high quality images of the carotid artery with a 2 cm depth and Doppler was activated to simultaneously measure blood velocity. Measurements were recorded for approximately 10 seconds using Camtasia screen capture software (Camtasia Studio 8, TechSmith, Okemos, MI). The right CCA was used for analysis of FCWI, using a custom-designed waveform analysis software developed by co-author Mynard in Matlab (The MathWorks Inc, Natick, MA, USA). Firstly, the arterial diameter waveform was measured across 3-5 cardiac cycles. Automated edge-detection was used to track the artery wall across the cardiac cycle to derive diameter waveforms from the B-mode image. Velocity waveforms were then calculated from the Doppler velocity, utilizing upper and lower envelope tracing to calculate the point-wise intensity-weighted spectral average to obtain a mean velocity waveform. Finally, the diameter and velocity waveforms were used to estimate wave speed and determine wave intensity in W/m^2^/s^2^. FCWI data were log-transformed for statistical analysis.

### Other Measures Assessed During the 28 Year Recall

At the 28-year follow-up appointment which included carotid ultrasound measurement of FCWI described above, participants had a variety of other measurements collected. Participants were seated in a cool, quiet room for 10-minutes, after which an automated blood pressure monitoring device (Carescape V100 Dinamap; General Electric Healthcare) took one measurement every 2 minutes for 10 consecutive minutes. Height was measured using a stadiometer with shoes removed, body weight was measured on seated scales with minimal clothing. Height and weight were used to calculate BMI (kg/m^2^). Participants were then placed supine on an assessment bed for 10 minutes, prior to the carotid artery ultrasound procedure being performed (described above). Following the carotid ultrasound assessment, a fasting venous blood sample was collected to test for glucose, insulin and lipids. Fasting glucose and insulin were used to calculate the homeostatic model assessment for insulin resistance (HOMA-IR) for each participant using the formula: glucose*insulin/22.5 (18). Participants completed an online questionnaire related to physical activity, time spent sitting, alcohol consumption and smoking habits.

### Statistical Analysis

Regression analysis was used (StataCorp, College Station, TX) to test for differences in FCWI for each risk factor trajectory (BMI, SBP, FBG) separately for sex. A secondary cross-section analysis tested whether variables measured at the 28-yr follow-up were also associated with risk factor trajectory group membership. These included: BMI, blood pressure, resting heart rate, low-density lipoprotein (LDL), high-density lipoprotein (HDL), HOMA-IR, self-reported physical activity (MET mins/wk), self-reported time spent sitting (mins/wk), alcohol consumption and frequency and smoking habits.

To test for cross-sectional differences in BMI, FBG, SBP and FCWI at age 28-years between males and females, independent samples *t*-tests or Wilcoxon Rank-Sum tests were used for data with normal or skewed distribution respectively.

## Results

Of the 1,997 participants invited to attend the 28-year follow-up appointment, 828 participants (n= 412 male, n= 416 female) accepted the invitation. Trajectories for BMI, SBP and FBG measured between ages 8-20 years were developed using the 828 participants that attended the 28-year follow-up and 565 received an ultrasound of the common carotid artery. Due to the high-quality images required to obtain accurate and valid analysis of FCWI, data for 402 participants (n=206 male, n=196 female) was of sufficient quality to undergo FCWI analysis and used for statistical analysis, see Figure 1. The mean age of participants at the 28-year follow-up appointment was 28.3±0.5 years and 28.3±0.6 years for males and females respectively.

**Figure 1.**
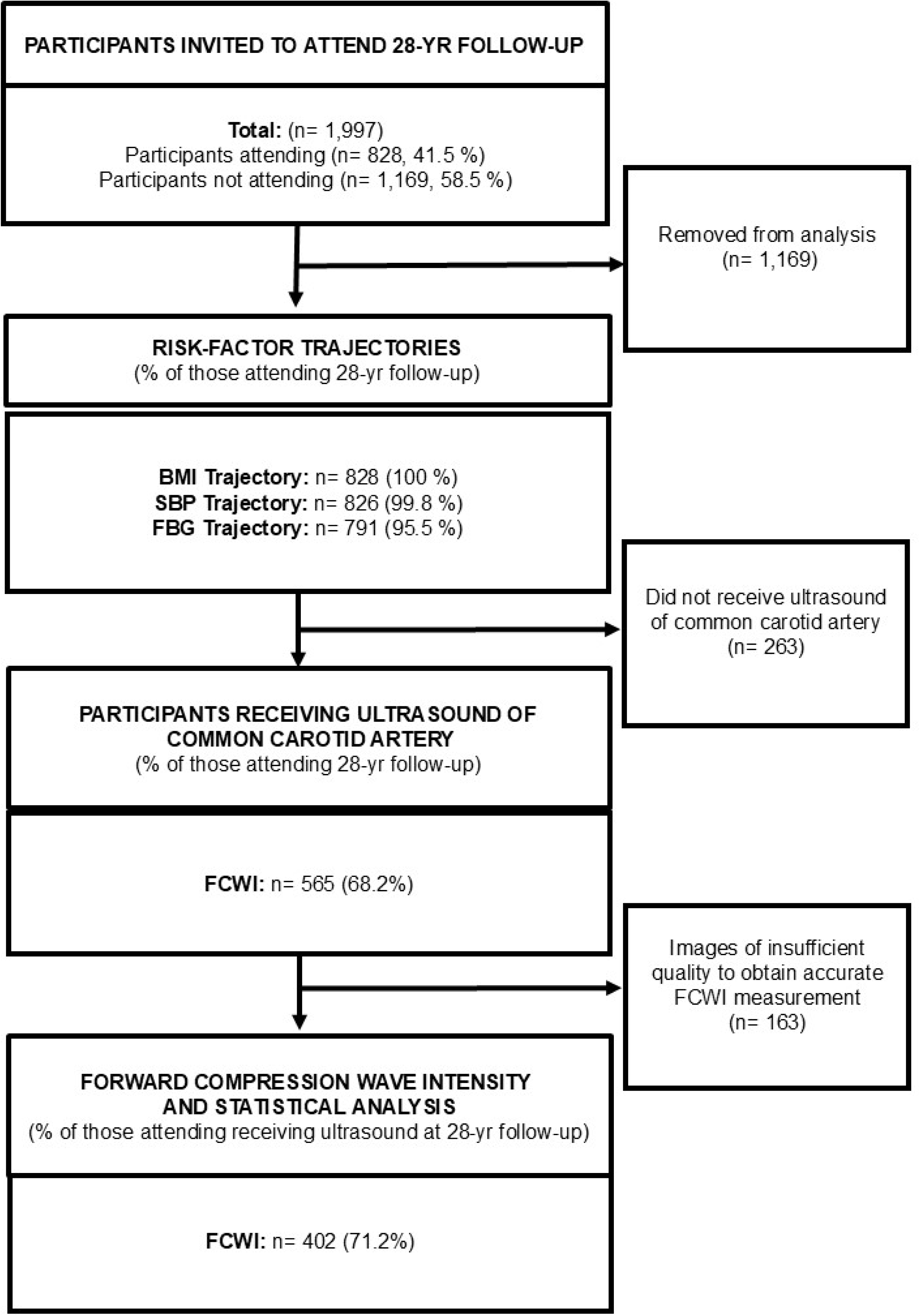
Consort Diagram indicating the number and percentages of participants contacted and assessed in the study. Body mass index (BMI), Systolic Blood Pressure (SBP), Fasting Blood Glucose (FBG), Forward Compression Wave Intensity (FCWI).

### Risk-Factor Trajectories

In males, four distinct trajectory groups were identified for BMI (see Figure 2 panel A), three trajectory groups were identified for SBP (see Figure 3 Panel A), and two trajectory groups were identified for FBG (see Figure 4 Panel A). Of the 828 participants that were included in the risk factor trajectory development, 2 female participants were not included in the SBP trajectory due to insufficient data (i.e. not having at least 3 timepoints available), and 37 participants (M=19, F=18) were not included in the glucose trajectory for the same reason. For all trajectories, group membership at age 8 years was predictive of membership throughout all subsequent follow-ups (i.e. no crossing-over of group trajectory patterns). Cross-sectional characteristics of male participants at the 28-year follow-up, separated for BMI trajectory groups can be found in Table 1, and for blood pressure and glucose trajectories can be found in Supplementary Table 1 and 2 respectively.

**Figure 2.**
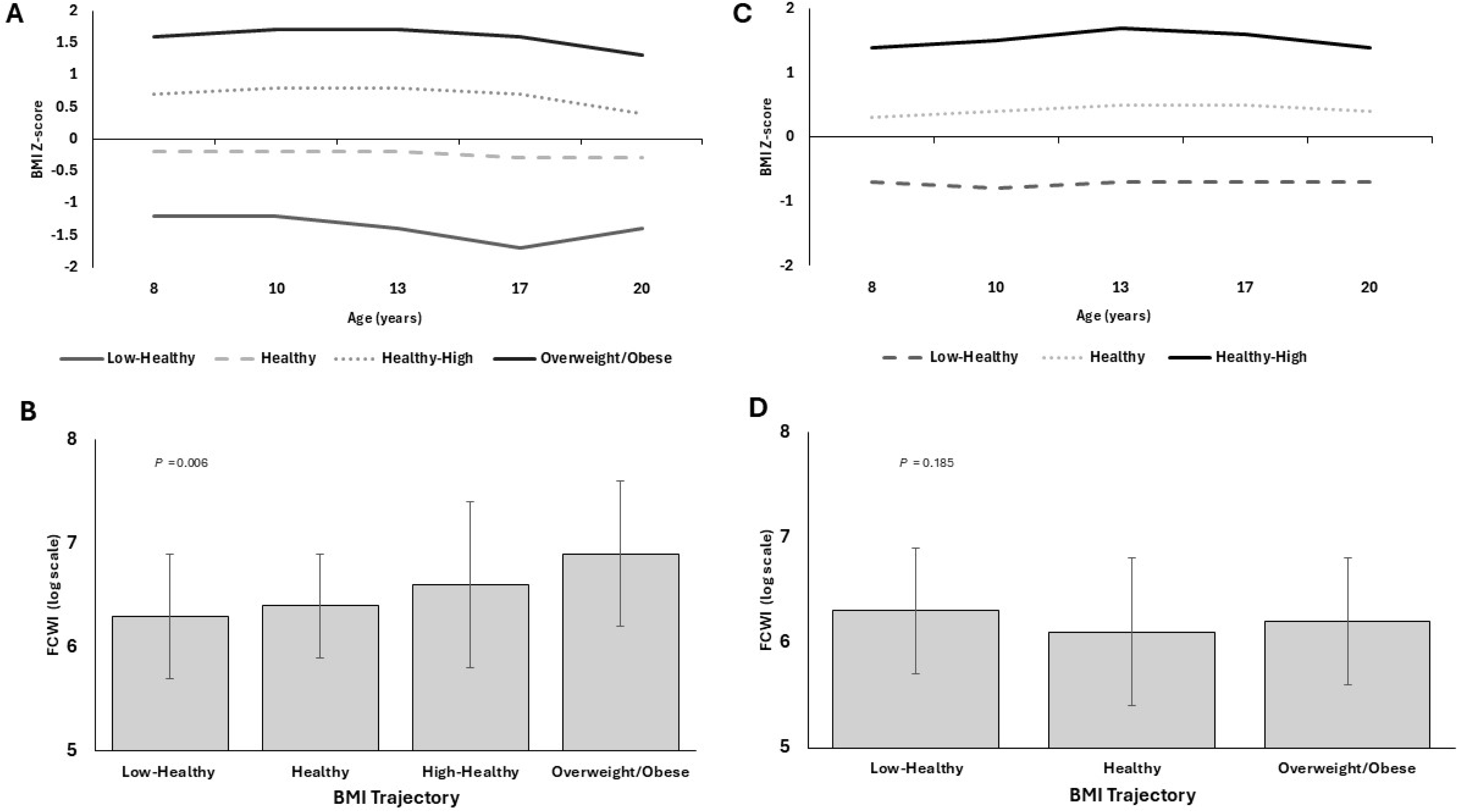
Body mass index (BMI Z-score) trajectories in male (Panel A) and female (Panel B) participants from ages 8-20 years and log-transformed forward compression wave intensity (FCWI) measured at age 28 years (mean±SD) for each BMI trajectory in males (Panel C) and females (Panel D). \**P*= derived from regression analysis to test for differences in FCWI between trajectory groups.

**Figure 3.**
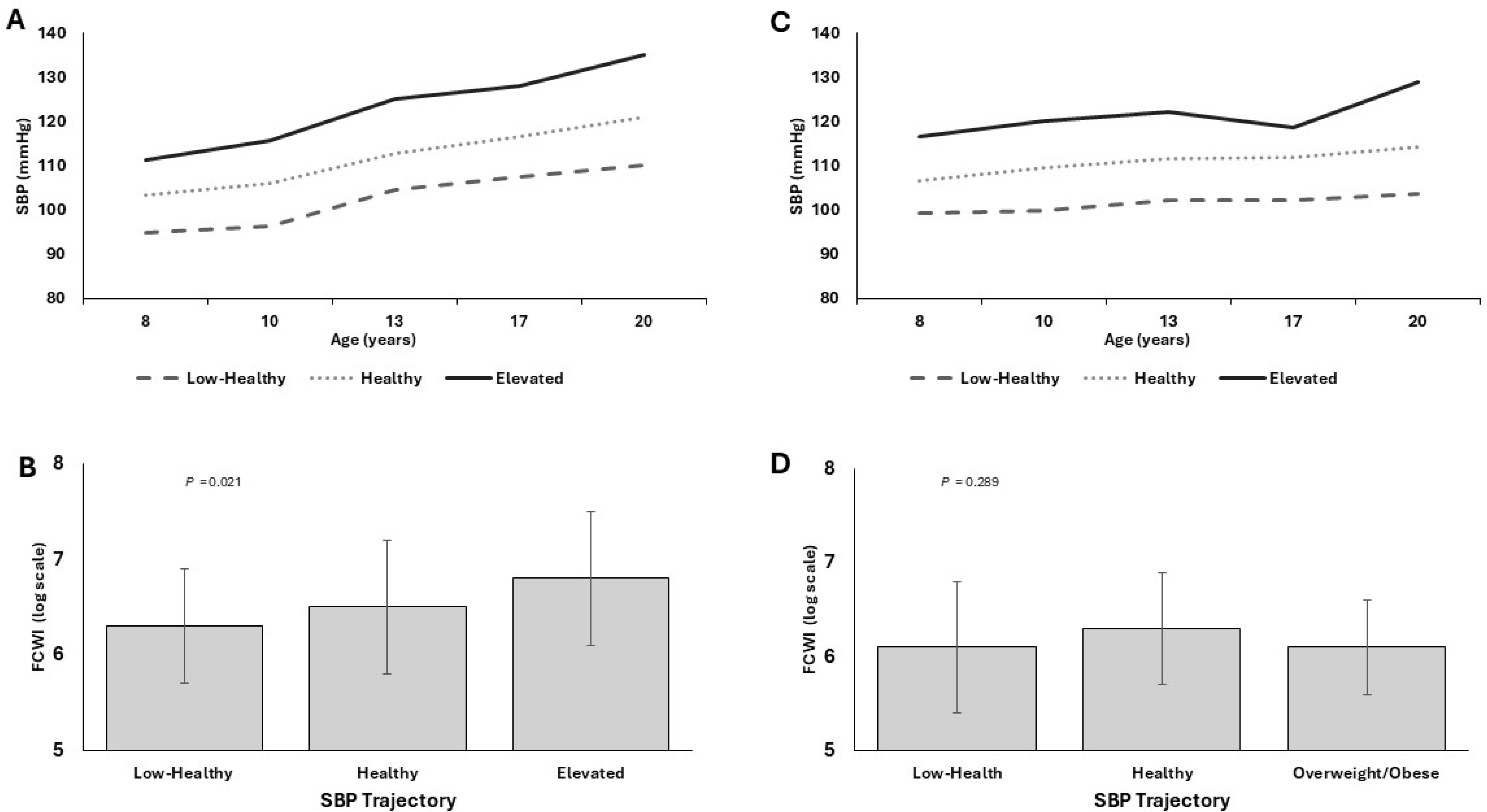
Systolic blood pressure (SBP) trajectories in male (Panel A) and female (Panel B) participants from ages 8-20 years and log-transformed forward compression wave intensity (FCWI) measured at age 28 years (mean±SD) for each SBP trajectory in males (Panel C) and females (Panel D). \**P*= derived from regression analysis to test for differences in FCWI between trajectory groups.

**Figure 4.**
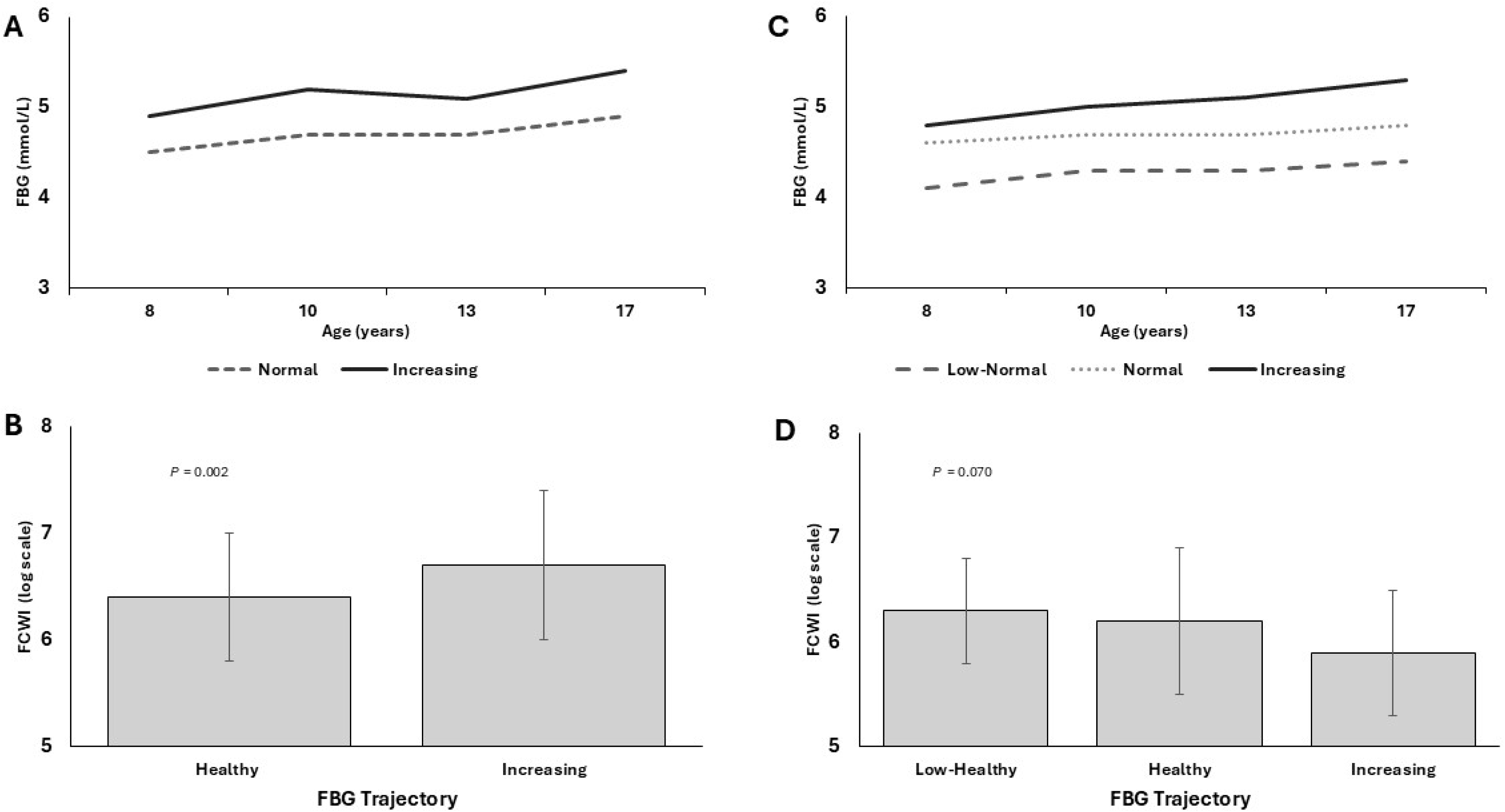
Fasting blood glucose (FBG) trajectories in male (Panel A) and female (Panel B) participants from ages 8-20 years and log-transformed forward compression wave intensity (FCWI) measured at age 28 years (mean±SD) for each FBG trajectory in males (Panel C) and females (Panel D). \**P*= derived from regression analysis to test for differences in FCWI between trajectory groups.

**Table 1.**
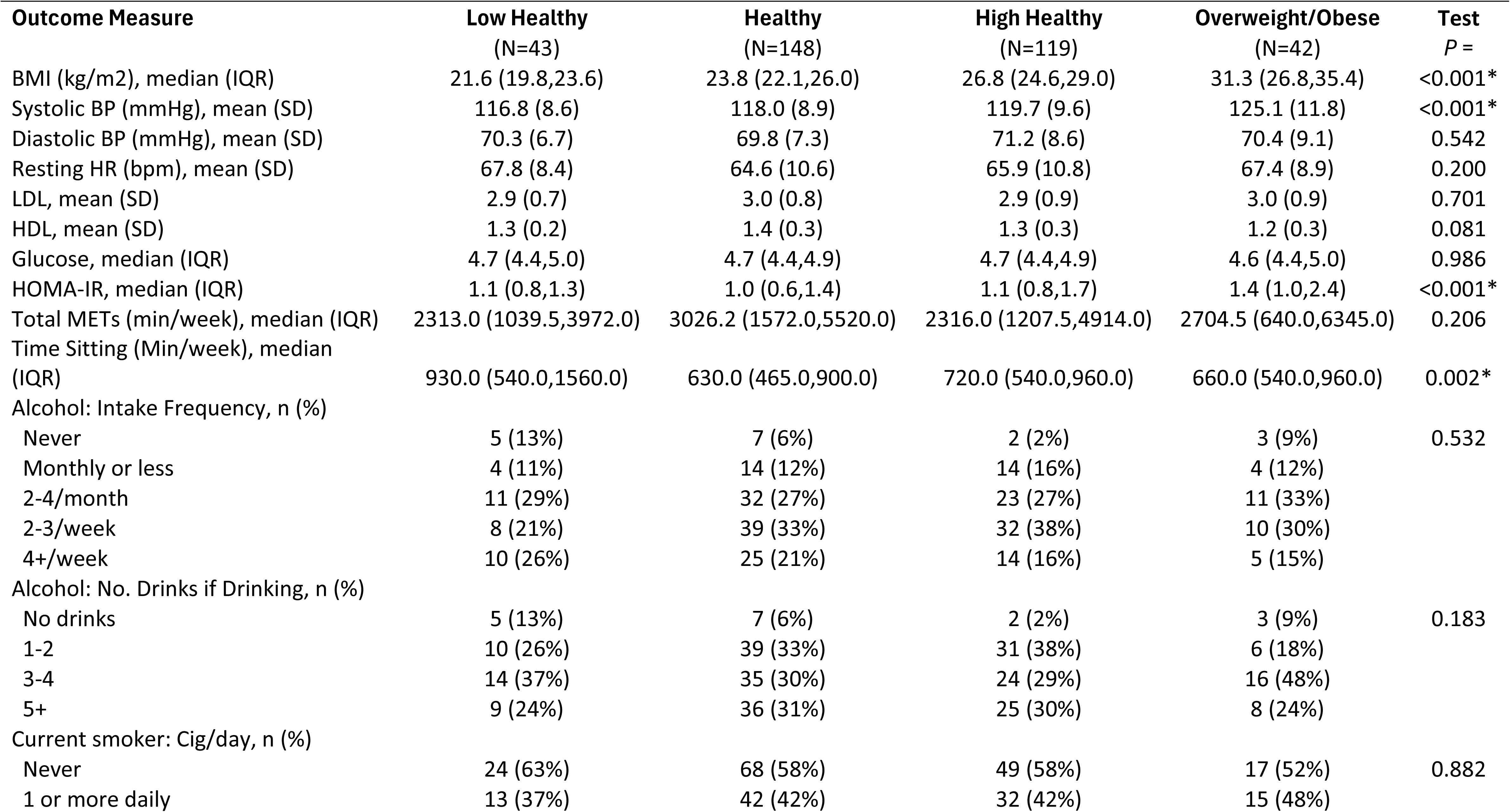
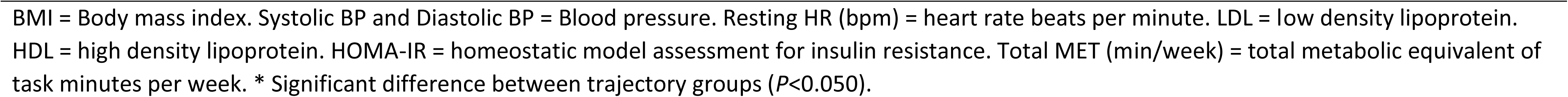
Characteristics of male participants at age 28-years based on body mass index trajectories from ages 8-20 years.

| Outcome Measure | Low Healthy<br>(N=43) | Healthy<br>(N=148) | High Healthy<br>(N=119) | Overweight/Obese<br>(N=42) | Test<br>P = |
| --- | --- | --- | --- | --- | --- |
| BMI (kg/m<sup>2</sup>), median (IQR) | 21.6 (19.8,23.6) | 23.8 (22.1,26.0) | 26.8 (24.6,29.0) | 31.3 (26.8,35.4) | <0.001* |
| Systolic BP (mmHg), mean (SD) | 116.8 (8.6) | 118.0 (8.9) | 119.7 (9.6) | 125.1 (11.8) | <0.001* |
| Diastolic BP (mmHg), mean (SD) | 70.3 (6.7) | 69.8 (7.3) | 71.2 (8.6) | 70.4 (9.1) | 0.542 |
| Resting HR (bpm), mean (SD) | 67.8 (8.4) | 64.6 (10.6) | 65.9 (10.8) | 67.4 (8.9) | 0.200 |
| LDL, mean (SD) | 2.9 (0.7) | 3.0 (0.8) | 2.9 (0.9) | 3.0 (0.9) | 0.701 |
| HDL, mean (SD) | 1.3 (0.2) | 1.4 (0.3) | 1.3 (0.3) | 1.2 (0.3) | 0.081 |
| Glucose, median (IQR) | 4.7 (4.4,5.0) | 4.7 (4.4,4.9) | 4.7 (4.4,4.9) | 4.6 (4.4,5.0) | 0.986 |
| HOMA-IR, median (IQR) | 1.1 (0.8,1.3) | 1.0 (0.6,1.4) | 1.1 (0.8,1.7) | 1.4 (1.0,2.4) | <0.001* |
| Total METs (min/week), median (IQR) | 2313.0 (1039.5,3972.0) | 3026.2 (1572.0,5520.0) | 2316.0 (1207.5,4914.0) | 2704.5 (640.0,6345.0) | 0.206 |
| Time Sitting (Min/week), median (IQR) | 930.0 (540.0,1560.0) | 630.0 (465.0,900.0) | 720.0 (540.0,960.0) | 660.0 (540.0,960.0) | 0.002* |
| Alcohol: Intake Frequency, n (%) |  |  |  |  |  |
| Never | 5 (13%) | 7 (6%) | 2 (2%) | 3 (9%) | 0.532 |
| Monthly or less | 4 (11%) | 14 (12%) | 14 (16%) | 4 (12%) |  |
| 2-4/month | 11 (29%) | 32 (27%) | 23 (27%) | 11 (33%) |  |
| 2-3/week | 8 (21%) | 39 (33%) | 32 (38%) | 10 (30%) |  |
| 4+/week | 10 (26%) | 25 (21%) | 14 (16%) | 5 (15%) |  |
| Alcohol: No. Drinks if Drinking, n (%) |  |  |  |  |  |
| No drinks | 5 (13%) | 7 (6%) | 2 (2%) | 3 (9%) | 0.183 |
| 1-2 | 10 (26%) | 39 (33%) | 31 (38%) | 6 (18%) |  |
| 3-4 | 14 (37%) | 35 (30%) | 24 (29%) | 16 (48%) |  |
| 5+ | 9 (24%) | 36 (31%) | 25 (30%) | 8 (24%) |  |
| Current smoker: Cig/day, n (%) |  |  |  |  |  |
| Never | 24 (63%) | 68 (58%) | 49 (58%) | 17 (52%) | 0.882 |
| 1 or more daily | 13 (37%) | 42 (42%) | 32 (42%) | 15 (48%) |  |

BMI = Body mass index. Systolic BP and Diastolic BP = Blood pressure. Resting HR (bpm) = heart rate beats per minute. LDL = low density lipoprotein. HDL = high density lipoprotein. HOMA-IR = homeostatic model assessment for insulin resistance. Total MET (min/week) = total metabolic equivalent of task minutes per week. \* Significant difference between trajectory groups ( $P < 0.050$ ).

In females, three distinct trajectory groups were identified for BMI (see Figure 3 Panel B), SBP (Figure 4 Panel B) and FBG (see Figure 5 Panel B). None of the trajectory patterns crossed paths, suggesting that risk factors measured at age 8 years were indicative of membership throughout childhood and adolescence up to 20-years of age. Cross-sectional characteristics of female participants at the 28-year follow-up, separated for BMI trajectory groups can be found in Table 2, and for blood pressure and glucose trajectories can be found in Supplementary Table 3 and 4 respectively.

**Table 2.**
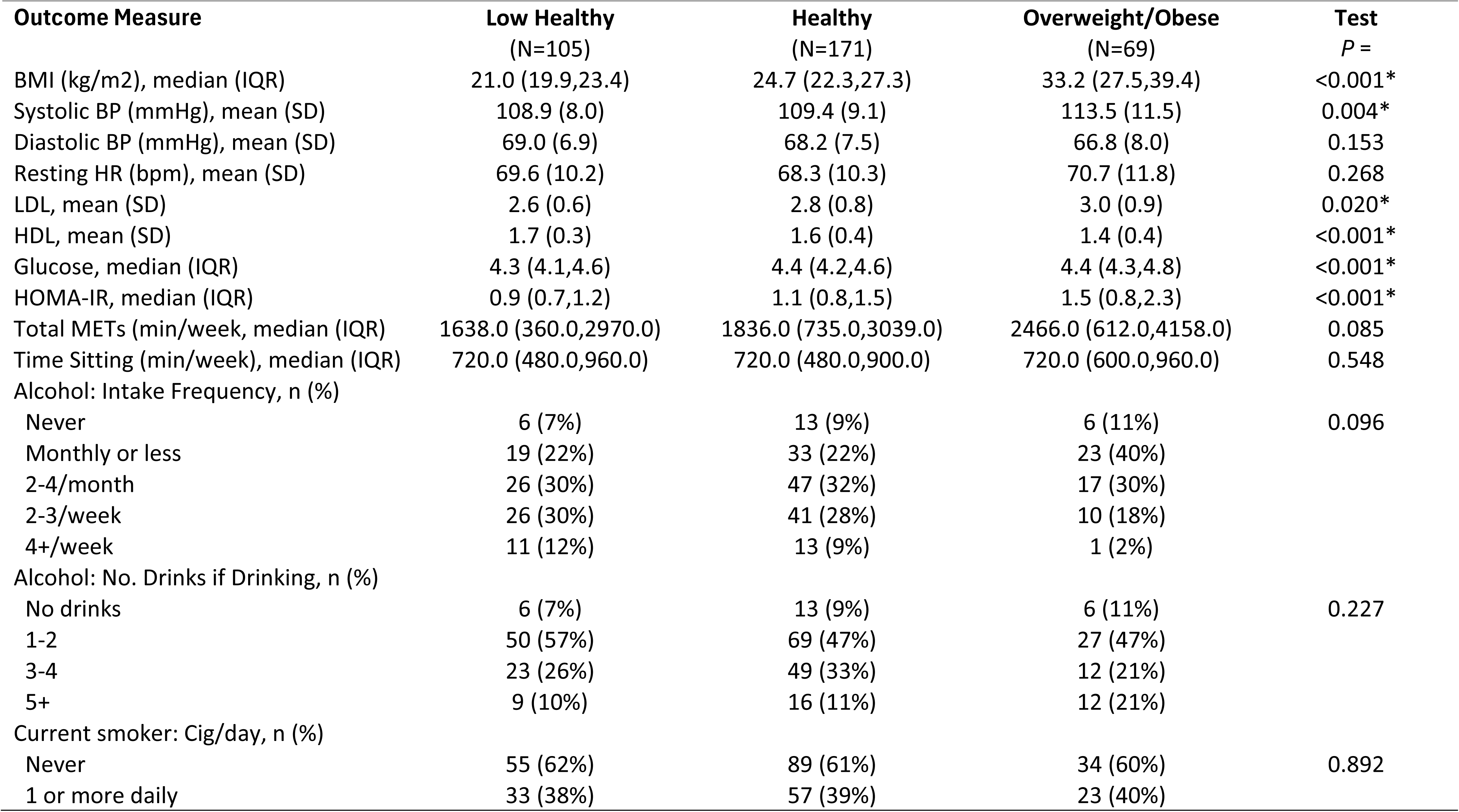
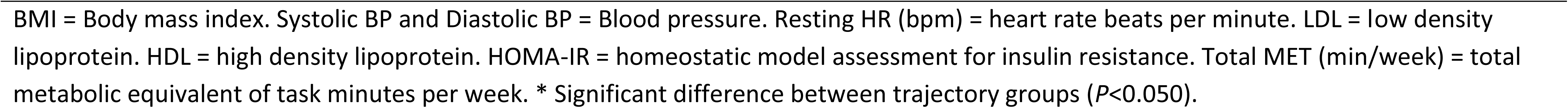
Characteristics of female participants at age 28-years based on body mass index trajectories from ages 8-20 years.

| **Outcome Measure** | **Low Healthy**<br>**(N=105)** | **Healthy**<br>**(N=171)** | **Overweight/Obese**<br>**(N=69)** | **Test**<br>**P =** |
| --- | --- | --- | --- | --- |
| BMI (kg/m<sup>2</sup>), median (IQR) | 21.0 (19.9,23.4) | 24.7 (22.3,27.3) | 33.2 (27.5,39.4) | <0.001* |
| Systolic BP (mmHg), mean (SD) | 108.9 (8.0) | 109.4 (9.1) | 113.5 (11.5) | 0.004* |
| Diastolic BP (mmHg), mean (SD) | 69.0 (6.9) | 68.2 (7.5) | 66.8 (8.0) | 0.153 |
| Resting HR (bpm), mean (SD) | 69.6 (10.2) | 68.3 (10.3) | 70.7 (11.8) | 0.268 |
| LDL, mean (SD) | 2.6 (0.6) | 2.8 (0.8) | 3.0 (0.9) | 0.020* |
| HDL, mean (SD) | 1.7 (0.3) | 1.6 (0.4) | 1.4 (0.4) | <0.001* |
| Glucose, median (IQR) | 4.3 (4.1,4.6) | 4.4 (4.2,4.6) | 4.4 (4.3,4.8) | <0.001* |
| HOMA-IR, median (IQR) | 0.9 (0.7,1.2) | 1.1 (0.8,1.5) | 1.5 (0.8,2.3) | <0.001* |
| Total METs (min/week, median (IQR) | 1638.0 (360.0,2970.0) | 1836.0 (735.0,3039.0) | 2466.0 (612.0,4158.0) | 0.085 |
| Time Sitting (min/week), median (IQR) | 720.0 (480.0,960.0) | 720.0 (480.0,900.0) | 720.0 (600.0,960.0) | 0.548 |
| Alcohol: Intake Frequency, n (%) |  |  |  |  |
| Never | 6 (7%) | 13 (9%) | 6 (11%) | 0.096 |
| Monthly or less | 19 (22%) | 33 (22%) | 23 (40%) |  |
| 2-4/month | 26 (30%) | 47 (32%) | 17 (30%) |  |
| 2-3/week | 26 (30%) | 41 (28%) | 10 (18%) |  |
| 4+/week | 11 (12%) | 13 (9%) | 1 (2%) |  |
| Alcohol: No. Drinks if Drinking, n (%) |  |  |  |  |
| No drinks | 6 (7%) | 13 (9%) | 6 (11%) | 0.227 |
| 1-2 | 50 (57%) | 69 (47%) | 27 (47%) |  |
| 3-4 | 23 (26%) | 49 (33%) | 12 (21%) |  |
| 5+ | 9 (10%) | 16 (11%) | 12 (21%) |  |
| Current smoker: Cig/day, n (%) |  |  |  |  |
| Never | 55 (62%) | 89 (61%) | 34 (60%) | 0.892 |
| 1 or more daily | 33 (38%) | 57 (39%) | 23 (40%) |  |

BMI = Body mass index. Systolic BP and Diastolic BP = Blood pressure. Resting HR (bpm) = heart rate beats per minute. LDL = low density lipoprotein. HDL = high density lipoprotein. HOMA-IR = homeostatic model assessment for insulin resistance. Total MET (min/week) = total metabolic equivalent of task minutes per week. \* Significant difference between trajectory groups ( $P < 0.050$ ).

### Impact of Risk-Factor Trajectories on Forward Compression Wave Intensity and Contemporaneous Risk Factors at age 28-years

For the BMI trajectory in males, there were significant differences in FCWI at age 28 years going from the lowest BMI (Low-Healthy) to highest BMI (Overweight/Obese), *P*=0.006, see Figure 3 Panel C. The overweight/obese trajectory group also had the highest HOMA-IR (*P*<0.001) at age 28 years, whereas the low-healthy BMI trajectory self-reported the greatest amount of sitting time at age 28 years (*P*=0.002), see Table 1.

For SBP trajectories in males, there were significant differences in FCWI at age 28 years going from the lowest SBP (Low-Healthy) to highest SBP (Elevated), *P*=0.021, see Figure 4 Panel C. The elevated SBP trajectory group also had the highest HOMA-IR (*P*=0.004) at age 28 years, see Supplementary Table 1.

In males, the highest FBG trajectory (Increasing) had significantly greater FCWI at age 28 years than the Healthy FBG trajectory (*P*=0.002), see Figure 5 Panel C, and also had higher HOMA-IR (*P*=0.045), see Supplementary Table 2.

In female participants, no significant differences were found in FCWI at age 28 years between any of the 3 trajectory groups: BMI (*P*=0.185, Figure 2 Panel D), SBP (*P*=0.289, Figure 3 Panel D) or FBG (*P*=0.070, Figure 4 Panel D).

### Relationship Between Variables Collected at Age 28-years

For contemporaneous risk factors measured at age 28-years in females (see Table 2), the “overweight” BMI trajectory had the highest BMI (*P<*0.001) and SBP (*P*=0.004), HOMA-IR (*P<*0.001) and LDL (*P*=0.020), and the lowest HDL (*P<*0.001). The “elevated” SBP trajectory had the highest BMI (*P<*0.001), SBP (*P<*0.001), diastolic BP (*P<*0.001), FBG (*P*=0.006) and HOMA-IR (*P*=0.003), see Supplementary Table 3. The Increasing FBG trajectory had the highest BMI (*P<*0.001), HDL (*P*=0.036), FBG (*P<*0.001) and HOMA-IR (*P*<0.001), see Supplementary Table 4.

### Differences Between Males and Females at age 28-years

There were significant differences between males and females for BMI, SBP, FBG and FCWI measured at age 28 years in the 402 participants that received FCWI analysis (all *P*<0.001), with males having higher values for each outcome compared to females (see Table 3).

**Table 3.**
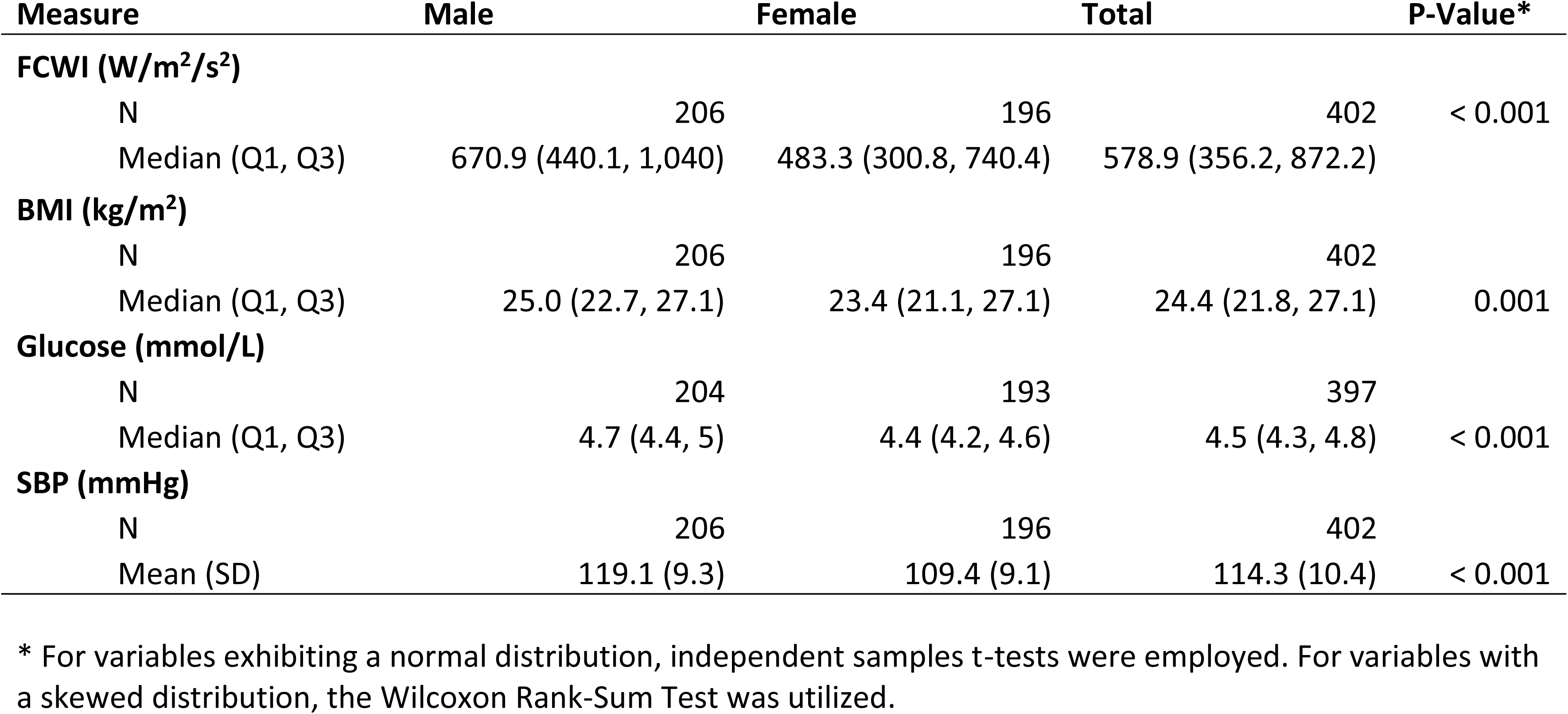
Sex differences in forward compression wave intensity (FCWI), body mass index, systolic blood pressure and fasting blood glucose at age 28-years in participants receiving FCWI analysis.

## Discussion

In this cohort study, we found that risk-factor trajectories derived from repeat measurements of body mass index, systolic blood pressure and fasting blood glucose from 8-20 years of age, were associated with greater forward compression wave intensity in the common carotid artery in males at age 28 years. Risk-factor trajectory membership was consistent throughout childhood and adolescence into adulthood in both males and females, suggesting that early-detection of those who may be at risk later in life, is possible in childhood. As high FCWI is an independent predictor of cognitive decline in a middle-older aged cohort (4), these novel results support the rationale for early detection and intervention, particularly in males, based on data collected at 28-years of age.

Wave intensity analysis of the carotid artery in humans is relatively novel, and utilizes pressure and velocity data to derive a measure of energy transfer per unit area, in the direction of the cerebral microcirculation (8, 19). It has been highlighted that, FCWI, which occurs in early systole, may be an important stimulus leading to mechanical damage of the cerebral microcirculation and associated neurons (4, 6). In the Whitehall-II study, which included >3,000 participants over a 10-year follow-up, individuals with the highest FCWI in middle-older age had the greatest rate of cognitive decline by age ∼60 years, and this was independent of other risk factors (4). In contrast to the Whitehall II study which began when participants were in mid-life, our study captured risk-factor data from childhood (age 8 years) to early adulthood (age 20 years), enabling assessment of antecedents that may be related to elevated FCWI into adulthood (i.e. age 28 years). The significant relationship between FCWI at age 28 years and BMI, SBP and FBG trajectories in males, which tracked well from childhood to adulthood in our cohort, complements the findings of the Whitehall study, suggesting that targeted prevention and risk factor modification strategies should commence early in life (13, 20, 21). Whilst currently speculative, it is plausible that when participants of the Raine cohort study age and exhibit overt symptoms or established disease manifests, future studies in this cohort may find that FCWI at age 28 years is predictive of cognitive decline, and possibly more so in males. In support of this, one cohort study found that childhood risk factor trajectories including elevated systolic blood pressure and blood lipids were negatively associated with cognitive performance in mid-life (22); whilst others have found associations between risk factors in childhood and established cardiovascular disease (CVD) in adulthood (23, 24). A study including >37,000 participants suggests that a single BP measurement at age 7 years of age was predictive of premature CVD mortality (median follow-up age of 54 years), with stronger associations in males than females (25).

The lack of association between risk factor trajectories and FCWI in females may be suggestive of a protective effect of being female, at least at this early life stage, potentially due to differences in the severity of these risk factors compared to males. Indeed, BMI, SBP, FBG and FCWI were all significantly lower in females compared to males in our cohort. The age of first occurrence of CVD is generally younger in males compared to females (26). In pre-menopausal females, the rates of cognitive decline, Alzheimer’s disease and CVD are lower than in age-matched males (27, 28), and the lower BMI, SBP and FCWI in females in our cohort at age 28 years may provide additional mechanistic evidence to support this phenomenon. Furthermore, evidence suggests that BP is generally lower in females compared to males until approximately 60 years of age, at which point females reach or may exceed BP observed in males (15, 29). It is possible that differences in FCWI between males and females also follow this age-related pattern, and may help explain why FCWI measurements were not associated with trajectory group membership in the pre-menopausal females in this study.

Approximately 25% of children and adolescents in Australia are classed as overweight or obese (30), and these rates are similar to other westernized countries (31), including the United States (32) and England (33). This indicates that a significant proportion of the growing populations in these countries would benefit greatly from targeted early prevention. A study which combined data from cohort studies to develop trajectories of BMI, SBP and total cholesterol from childhood to adulthood, found these variables were significant predictors of adult CVD diagnosis and events at age ∼49 years (23). This study also found that childhood risk factors tracked well into adulthood, resulting in childhood data being independently predictive of adult CVD in a sub-analysis; despite only one timepoint often being available in childhood (23). Our data, in which the FCWI outcome measures were collected when participants were aged ∼28 years, does not yet allow for clinical endpoints to be assessed. However, our data which includes measurements from multiple time-points from childhood to early-adulthood, indicates that these risk factors track well across this 20-year follow-up period to age 28 years, and that they are associated with FCWI in males, which is a novel and important predictor of cognitive decline (4).

This study has several limitations. The FCWI analysis was only performed in a sub-sample of the cohort, although it does not appear that this negatively impacted statistical power, at least in males. Because FCWI was measured at an age significantly younger than that associated with overt symptoms and CVD manifestation, and we did not measure cognitive function, we are not able to make definitive associations between high FCWI, future cognitive decline (4) or cerebrovascular risk in our cohort. However, the Raine cohort is followed regularly, and tracking of disease diagnoses as the cohort ages holds the potential to shed light on these areas. A strength of this study is the number of regular follow-ups participants attended in childhood and adolescence, allowing risk-factor trajectory development, and inclusion as a predictive variable against FCWI. FCWI analysis is an important emerging research tool and the quality of the duplex ultrasound assessments and subsequent FCWI analysis were of a high quality. Future research should investigate the time course at which FCWI increases in females, and whether this tracks with an increase in risk factors such as increased blood pressure and/or BMI, or relevant life-stages such as peri- or post-menopause (26–28).

In summary, this study utilized risk factor data collected over a 12-year period when participants were aged between 8 and 20 years. We used this data to develop sex-specific risk factor trajectories, which indicated that BMI, SBP and FBG measured in childhood and adolescence tracked well into adulthood. We present novel evidence that risk factor trajectories in males were predictive of FCWI at age 28 years. This may be predictive of cerebral-related pathologies later in life. Indeed, we have shown that the greater mechanical assault to the cerebral vasculature which occurs with every heartbeat (i.e. ∼100,000 times per day), is present for decades prior to the age generally associated with overt symptom development. These findings suggest that risk factor modification should begin as early as childhood, as this may result in favorable adaptations that may persist into adulthood and reduce risk. We also provide evidence to suggest that the risk factor profile that may contribute to the development of CVD, follows a different pattern and chronology in females compared to males.

## Data Availability

The data that support the findings of this study are available from the corresponding author upon reasonable request.

## Acknowledgements

We gratefully acknowledge all Raine Study participants and their families for their continued participation in the study, as well as the Raine Study team for study co-ordination and data collection. We also thank the NHMRC and the Raine Medical Research Foundation for their support. The core management of the Raine Study is funded by The University of Western Australia, Curtin University, The Kids Research Institute Australia, Women and Infants Research Foundation, Edith Cowan University, Murdoch University, The University of Notre Dame Australia and the Western Australian Future Health Research and Innovation Fund (Grant ID WACSOSP2023-2024, WACSOSP2025/7).

